# Functionality-Informed Fine-Mapping Dissects Common Variant Contributions to Coronary Artery Disease and Identifies Causal Variants and Pathways

**DOI:** 10.64898/2026.04.01.26349823

**Authors:** Jacob Thoft Jacobsen, Peter Loof Riis Møller, Palle Duun Rohde

## Abstract

**Summary:** Genomics offer a powerful approach to identify causal mechanisms underlying coronary artery disease (CAD) risk, with implications for pathogenesis, personalized prevention strategies, and therapeutic target discovery.

Functionality-informed genome-wide fine mapping was performed using the Bayesian framework SBayesRC to estimate genetic contributions of 6.9 million common variants, based on GWAS summary statistics from over one million individuals of European ancestry. Causal candidate genes were prioritized in a 5kB flanking window within high-confidence local credible sets (LCSs). Their downstream biological influence was analyzed using protein-protein interaction networks and pathway enrichment analyses across three complimentary dimensions: molecular, cellular, and disease level.

Genetic modeling captured the highly polygenic architecture of CAD, estimating on average 34,000 variants to contribute to CAD risk, explaining 3.8% of total phenotypic variance. 36 high-confidence variants (PIP > 0.9) collectively explained 13.6% of genetic variance, while most variants demonstrated small individual effects but with substantial collective contributions. 17,150 variants were prioritized within 581 high-confidence LCSs, of which 195 were annotated to genes and 170 were implicated in downstream pathway analyses. The three most influential variants were mapped to *PHACTR1, APOE*, and *LPL*, explaining 2.49%, 1.59%, and 1.46% of genetic variance respectively. Pathway analyses revealed that genetic risk in CAD is driven by dysregulation of three interlinked biological processes: 1) lipoprotein function and cholesterol metabolism, 2) vascular homeostasis, and 3) cellular stress responses and inflammation.

These findings advance the causal understanding of CAD pathogenesis, supporting the transition from association-based to functionality-informed genomic approaches in cardiovascular genetics.

## Background

Coronary artery disease (CAD) remains a global health burden and persists as the leading cause of morbidity and mortality worldwide. In 2022, atherosclerotic cardiovascular disease caused 19.8 million deaths globally, representing approximately a third of all deaths. CAD, the lead killer, accounted for 7.5 million of these deaths [1]. The global prevalence of CAD has increased from 100 million in 1990 to more than 180 million in 2019. Thus, CAD is of increasingly global concern, even in high-income countries such as the USA and UK, in which strategies for disease management have seemed to fail in recent years [2].

Atherosclerosis is characterized as the progressive accumulation of lipid-rich and fibrous deposits (plaques) within the intima, the innermost layer of the arterial wall (Figure 1) [3]. It is a chronic condition developed throughout life by exposure to risk factors that contribute to accumulation of disease risk over many years and decades. Traditional risk factors include older age, male sex, family history of premature disease, tobacco use, hypertension, hypercholesterolemia, diabetes mellitus, unhealthy diet, obesity, physical inactivity, and high alcohol usage [4].

**Figure 1:**
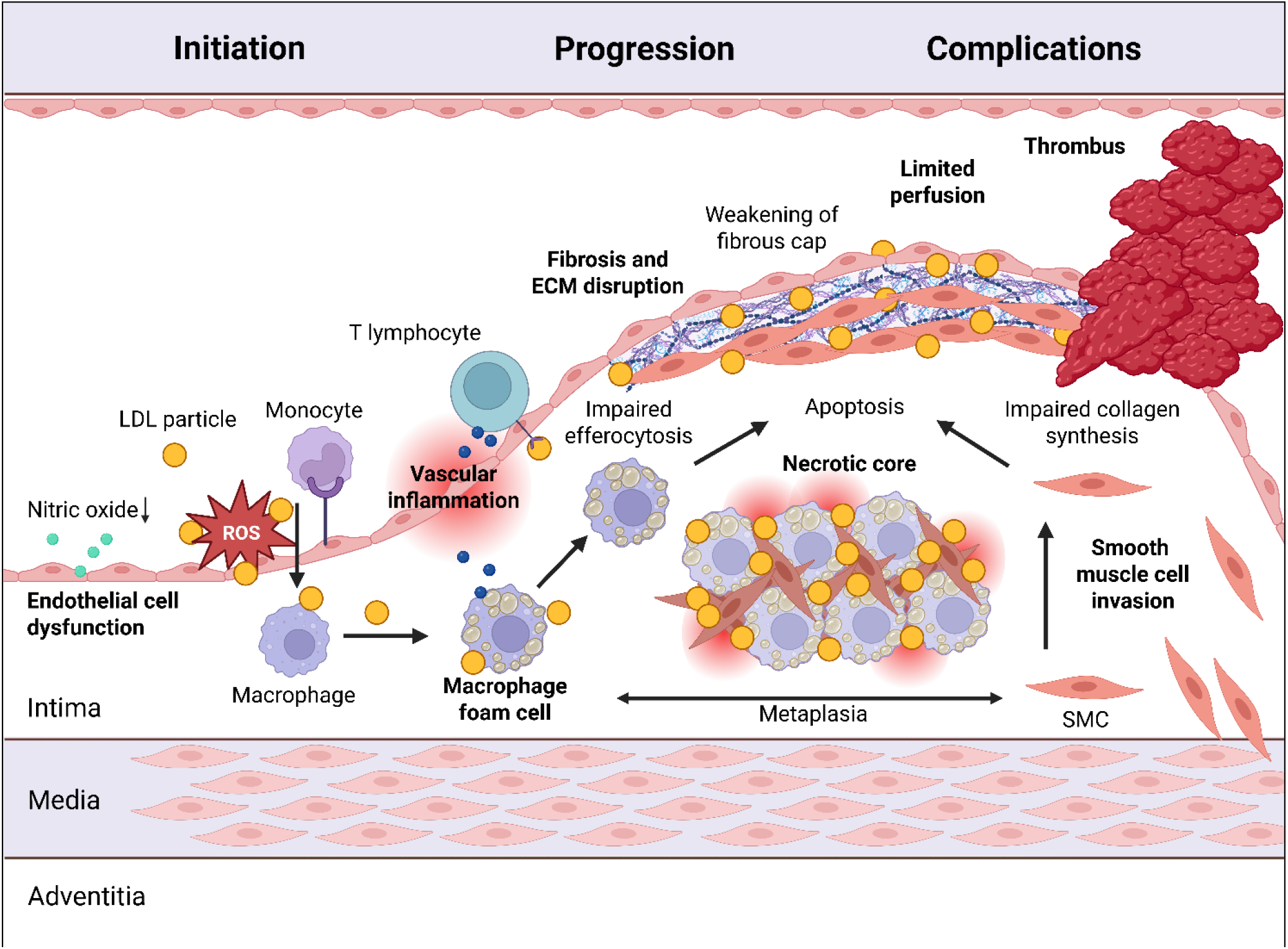
Pathogenesis of atherosclerosis. Atherosclerosis is characterized as the life-long accumulation of lipid rich and fibrous deposits within the tunica intima that forms plaques. Atherosclerosis is initialized by entrapment and accumulation of LDL particles in the intima. Here, oxidative and other modifications of the LDL particles give rise to proinflammatory and immugenic properties. Circulating monocytes are recruited to the beginning plaque where they differentiate into macrophages, internalize the LDL particles, and turn into macrophage foam cells with excessive proinflammatory functions. Byproducts from modified LDL particles may function as epitopes that activate the adaptive immune system. T lymphocytes enter the intima and orchestrate the microenvironment, modulating endothelial, immune, and smooth muscle cell function. Smooth muscle cells are recruited and invade the intima from the tunica media. Progression of the plaque continues as lipid-rich deposits, lipid-laden foam cells, and smooth muscle cells accumulate and form the bulk of the plaque. Recruited immune cells drive the chronic vascular inflammation. Smooth muscle cells may undergo metaplasia and shift into foam cells resembling those of macrophages. Macrophages and smooth muscle cells undergo apoptosis, while impaired efferocytosis leads to reduced clearance of cell debris, creating the necrotic lipid-rich core. Disturbed homeostasis in extracellular matrix deposition leads to further entrapment of lipids, reduced collagen synthesis, and increased enzymatic activity that degrade the fibrous cap overlying the core. The plaque may calcify, further weakening the fibrous cap. Complications of atherosclerosis arise when the plaque limits blood flow either due to stenosis or thrombotic occlusion. Flow-limiting stenosis happens when the plaque occludes 50-75% of the lumen, resulting in chronic reduced perfusion with stable angina pectoris as symptom. Thrombotic occlusion happens due to plaque rupture or erosion, resulting in acute coronary syndrome with unstable angina pectoris as symptom. LDL: low-density lipoprotein. ECM: extracellular matrix. Made in BioRender.

In recent decades, the risk profile has shifted, possibly reflecting better control of the traditional risk factors [5]. The disease now more frequently affects females, younger individuals, and ethnically diverse individuals [6]. Even for patients that have achieved their treatment goals of lifestyle changes and lowering of LDL levels, cardiovascular events still occur [3].

Research has thus expanded the search for new risk-contributing markers to improve disease management. Major advancements within the fields of omics technologies during the last decade provide promises in identification of useful novel molecular biomarkers. Genomics may provide substantial value in the future as part of a precision-medicine approach to enhance risk profiling and support individual-level management of CAD [7]. Heritability is estimated to account for 40-60% of the total phenotypic variance in CAD [8]. During the last decade, large-scale genome-wide association studies (GWAS) have identified approximately 400 genome-wide disease-associated genetic loci in CAD [9].

The majority of the identified risk loci are in intergenic genomic regions, suggesting they act on gene expression rather than protein level [10]. They predominantly influence physiological pathways related to lipid metabolism and vascular function. However, the exact mechanisms of the physiological effect for the majority of identified variants remain unsettled [7]. A key focus in recent studies as well as future work involves this link from association to physiological mechanism.Physiological understanding enables risk prevention and drug development and plays a major part in the precision-medicine approach [7].

Genomics may provide insights into causal mechanistic physiological functions that drive CAD risk. This study therefore aims to dissect the genetic contribution in CAD utilizing GWAS summary statistics by 1) modeling genetic effects and prioritizing candidate variants for causal roles, 2) identifying influential genetic variants that individually contribute substantially to heritability, 3) analyzing their involvement in downstream biological pathways that drive genetic risk, and 4) contextualizing these findings within the existing evidence on CAD pathogenesis.

## Methods

### Functionality-informed Genome-wide Fine Mapping

Joint association analysis was performed using SBayesRC [11] integrated in the GCTB software (version 2.5.4) as part of the functionality-informed genome-wide fine mapping [12].

Baseline GWAS summary statistics were obtained from Aragam et al. [13]. The dataset included 20,073,070 genetic variants from 181,522 CAD cases among 1,165,690 total participants of predominantly European ancestry (> 95%). Rare variants (MAF < 0.01), insertions, deletions and multi-allelic sites were excluded to only include common autosomal single nucleotide polymorphisms (SNPs).

Linkage disequilibrium (LD) data and functional annotations (BaselineModel2.2) were obtained from Zheng et al. [11] as part of their SBayesRC GitHub resource. LD data covered 7,356,518 SNPs derived from 20,000 individuals of European Ancestry in the UK Biobank. Functional annotations consisted of SNP-level enrichment values for 8M SNPs in each of 96 categories of genomic positions or functions [14].

Prior to SBayesRC modeling, quality control steps were performed to align GWAS summary statistics and LD data using the SBayesRC R package. SNPs with non-consistent alleles, SNPs with large frequency mismatches (Freq_thresh = 0.2), SNPs outside sample size mean ± 3×SD (n_range = 3), and SNPs with variance explained inconsistencies (rate2pq =0.5) were excluded. Flipped alleles (3,269,494 SNPs) were properly handled and imputation of summary statistics were performed to match the SNP set present in LD data. This resulted in 6,948,252 common SNPs intersecting between GWAS summary and LD data.

The SBayesRC model was established by setting SNP-based heritability to a starting value of hsq = 0.5. Starting proportions in each mixture component was set to *π* = 0.99, 0.005, 0.003,0.001, and 0.001, assuming 99% of SNPs having no effect, 0.5% of SNPs having tiny effect, 0.3% of SNPs having small effect, 0.1% of SNPs having moderate effect, and 0.01% of SNPs having large effect. Starting component variance scaler was set to *γ* = 0, 0.001, 0.01, 0.1, and 1,assuming SNPs in each mixture component to explain from 0% to 1% of genetic variance, respectively.

Sampling of the posterior distributions was performed with MCMC algorithm over a total of 6000 iterations with 2000 iterations as burn-in. Final genetic effect sizes were estimated as the robust posterior mean across the 4000 post burn-in iterations. During modeling, SNP effect sizes below a threshold of 0.0014 were shrunk to 0 to only keep meaningful signals.

### Prioritization of Candidate Causal Genes

Candidate casual genes were prioritized by constructing local credible sets (LCSs) based on SBayesRC sampling of SNP-level posterior inclusion probability (PIP) and posterior SNP-based heritability enrichment probability (PEP). LCS were constructed using GCTB software (version 2.5.4) [12]. In short, SNP candidate pools were established within 591 LD blocks. In these, LCSs were constructed by selecting SNPs with a pairwise LD r^2^ > 0.5 with a lead PIP candidate (rsq 0.5). High confidence sets were established by filtering for an accumulated sum of PIP ≥ 0.9 (pip 0.9) and PEP ≥ 0.7 (pep 0.7).

High confidence LCSs were mapped to the nearest gene with a flanking window of 5Kb (flank 5000) using a gene map provided by the GCTB resource consisting of 63,192 positional gene annotations.

### Protein-protein Interaction Network and Cluster Analysis

Based on prioritized genes, protein-protein interaction networks were established and analyzed using the database for Homo Sapiens and software on the STRING website (https://string-db.org/).

All possible interaction evidence was integrated (textmining, experiments, databases, co-expression, neighborhood, gene fusion, and co-occurrence). To ensure only high-confidence interactions, a minimum required interaction score of 0.700 was applied. Disconnected proteins were excluded in the visualization of the protein network to remove visual noise.

Hub genes in the network with high degree of interactions (≥ 5) were identified as essential players with key biological functions in CAD pathogenesis. To identify biological categories within the protein-protein interaction network, k-means clustering was performed, representing groups of tightly interconnected proteins likely related to meaningful biological processes contributing to CAD risk.

### Pathway Enrichment Analysis

Pathway enrichment analysis of prioritized genes was performed across three complementary dimensions using the STRING software and integrated databases.

Molecular-level enrichment was assessed using the Reactome database [15] to assess whether the prioritized genes were overrepresented in specific biological pathways. Tissue-level enrichment was assessed using the TISSUES database [16] to provide insights into tissues that overexpress the prioritized genes. Disease-level enrichment was assessed using the DisGeNET database [17] to validate the associations of prioritized genes to disease-relevant phenotypes.

## Results

### Genetic Variance Explained in Mixture Components

An average of 34,468 (0.5% of total) SNPs were estimated to contribute with non-zero effects.SNP-based heritability was estimated to explain 3.8% of total phenotypic variance.

99.5% of SNPs were assigned to have zero genetic effect (mixture component 1). Among non-zero variants, the distribution of genetic effect followed the expected pattern for complex diseases. Most variants demonstrated small individual contributions but collectively explained substantial variance. SNPs with small effects (mixture component 3) were contributing strongest to heritability, explaining 41% of genetic variance despite representing only 13.6% of non-zero variants. In contrast, only 14 SNPs were on average assigned to having large individual effect (mixture component 5), collectively explaining 16% of genetic variance (Table 1).

**Table 1:** Average non-zero mixture component distributions and their contributions to genetic variance (Var_g_).

| Component | Genetic Effect | SNPs | $Var_g$ |
| --- | --- | --- | --- |
| 2 | Tiny | 29,552 (85.8%) | 26% |
| 3 | Small | 4698 (13.6%) | 41% |
| 4 | Moderate | 204 (0.6%) | 17% |
| 5 | Large | 14 (0.0%) | 16% |

### Estimations of Joint Genetic Effects and Causality

Across all 4000 post burn-in iterations, 1,495,206 (21.5% of total) SNPs demonstrated a non-zero posterior mean, representing possible candidates for causal genetic associations with CAD.

The distribution of effect sizes was heavily right-skewed with the majority of SNPs having extremely small effect sizes (median |β| = 2.4 × 10−5, |β|min = 1 × 10−6, |β|max = 0.36). The 95th percentile of absolute effect sizes was 2 × 10−4, illustrating the predominance of variants with minimal individual contribution. Of causal candidates, 99.98% SNPs had small effect (|*β*| < 0.01), 323 SNPs had moderate effect (|*β*| = 0.01 - 0.1), and 8 SNPs had large effect (|*β*| > 0.1), (Figure 2a).

**Figure 2:**
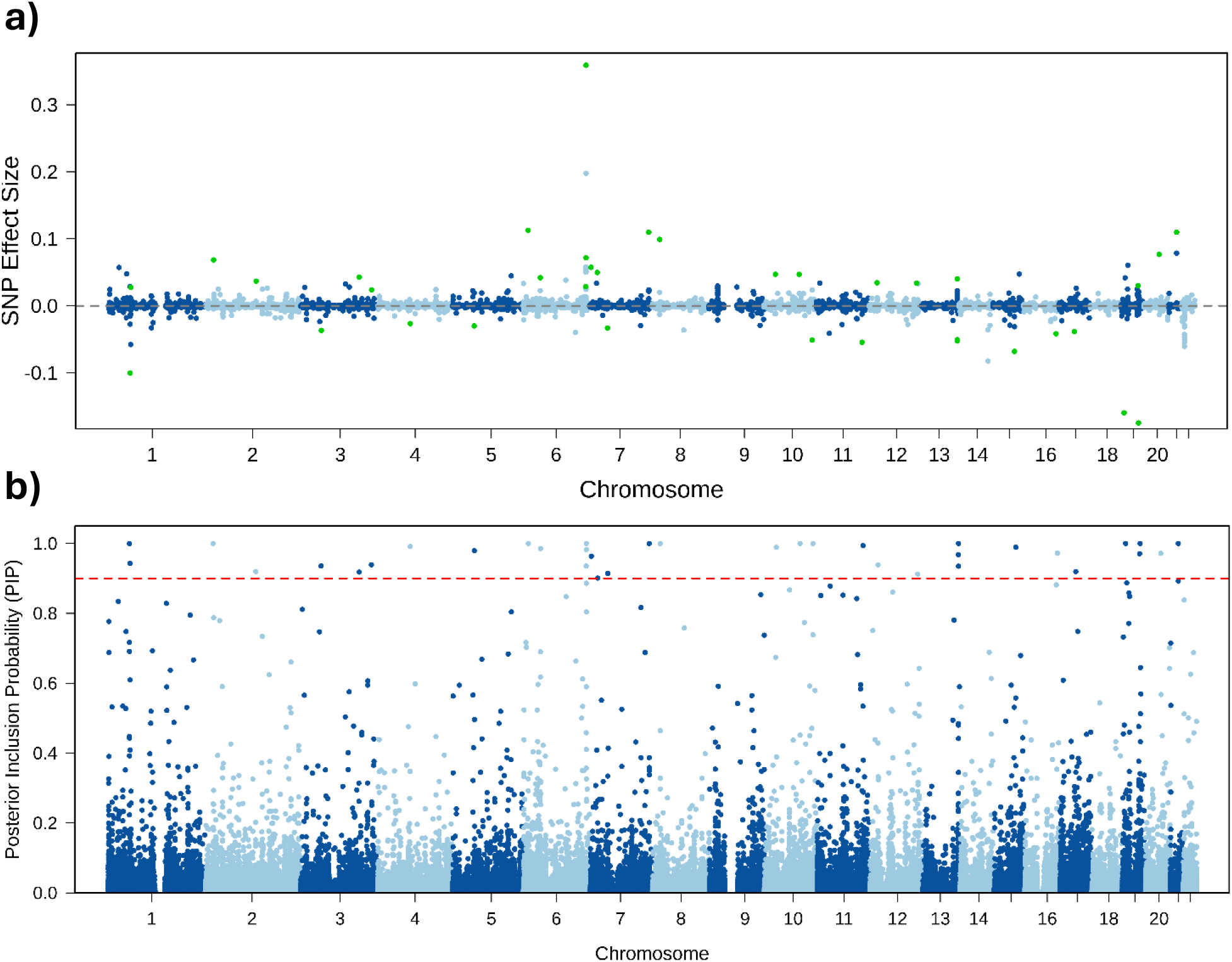
**a) Estimated joint effect size of genetic variants**. Genome-wide joint associations with CAD were modeled using SBayesRC based on GWAS summary statistics, LD data, and functional annotations. Joint effect sizes were estimated as the posterior mean of 4000 MCMC iterations. Non-zero effect SNPs estimated as causal candidates (n = 1.5M SNPs) are plotted with high-confidence SNPs (PIP > 0.9) highlighted in green. Joint effect modeling identified few genetic variants with large individual effects. In contrast, most variants were estimated to contribute very little to genetic variance individually, however explaining much of genetic variance collectively. **b) Estimated probabilities of causality based on PIP**. PIP estimations for genome-wide non-zero SNPs (n = 1.5M SNPs). SBayesRC modeling revealed very few high confidence SNPs (n = 36) with large individual genetic contributions. The majority of modeled SNPs were estimated as low confidence causal associations, individually contributing with minor genetic effect but collectively explaining large parts of genetic variance. The red line highlights the high-confidence PIP threshold of 0.9.

Modeling identified 36 very high confidence SNPs (PIP > 0.9) collectively explaining a total of 13.6% of genetic variance. In contrast, 99.8% of all non-zero SNPs were categorized as very low confidence SNPs (PIP < 0.1), collectively contributing to 60.6% of genetic variance (Figure 2b and Table 2).

**Table 2:** Causal confidence of non-zero variants and their contribution to genetic variance (Var_g_).

| Causal confidence (PIP) | SNPs | Mean $\beta$ | $Var_g$ |
| --- | --- | --- | --- |
| Very Low ($\leq 0.1$) | 99.8% | $5 \times 10^{-5}$ | 60.6% |
| Low (0.1 - 0.4) | 2826 | $2 \times 10^{-3}$ | 16.5% |
| Moderate (0.4 - 0.6) | 129 | $1 \times 10^{-2}$ | 4.0% |
| High (0.6 - 0.9) | 69 | $3 \times 10^{-2}$ | 5.3% |
| Very High ($> 0.9$) | 36 | $7 \times 10^{-2}$ | 13.6% |

### Genetic Variance Explained Across Allele Frequencies

The contribution to genetic variance across allele frequency bins (Table 3) demonstrated that approximately half of total genetic variance was explained by SNPs with frequencies < 0.25. Results demonstrated a tendency that low-frequency variants had greater effect sizes than more common variants (Figure 3), an evolutionary signature of natural selection.Analysis of effect sizes across allele frequencies demonstrated that both risk-increasing and protective causal candidates (PIP > 0.5) were present across the whole frequency range.

**Table 3:** Estimated contributions to genetic variance across allele frequencies (Var_g_).

| Allele Frequency | SNPs | Mean $\beta$ | $Var_g$ |
| --- | --- | --- | --- |
| Low (0.01 - 0.05) | 14.4% | $1.2 \times 10^{-4}$ | 12.5% |
| Moderate (0.05 - 0.1) | 9.6% | $8.3 \times 10^{-5}$ | 11.2% |
| Common (0.1 - 0.25) | 24.5% | $6.1 \times 10^{-5}$ | 23.9% |
| Very common (0.25 - 0.5) | 28.3% | $5.1 \times 10^{-5}$ | 31.7% |
| Extremely common ($> 0.5$) | 23.0% | $5.3 \times 10^{-5}$ | 20.8% |

**Figure 3:**
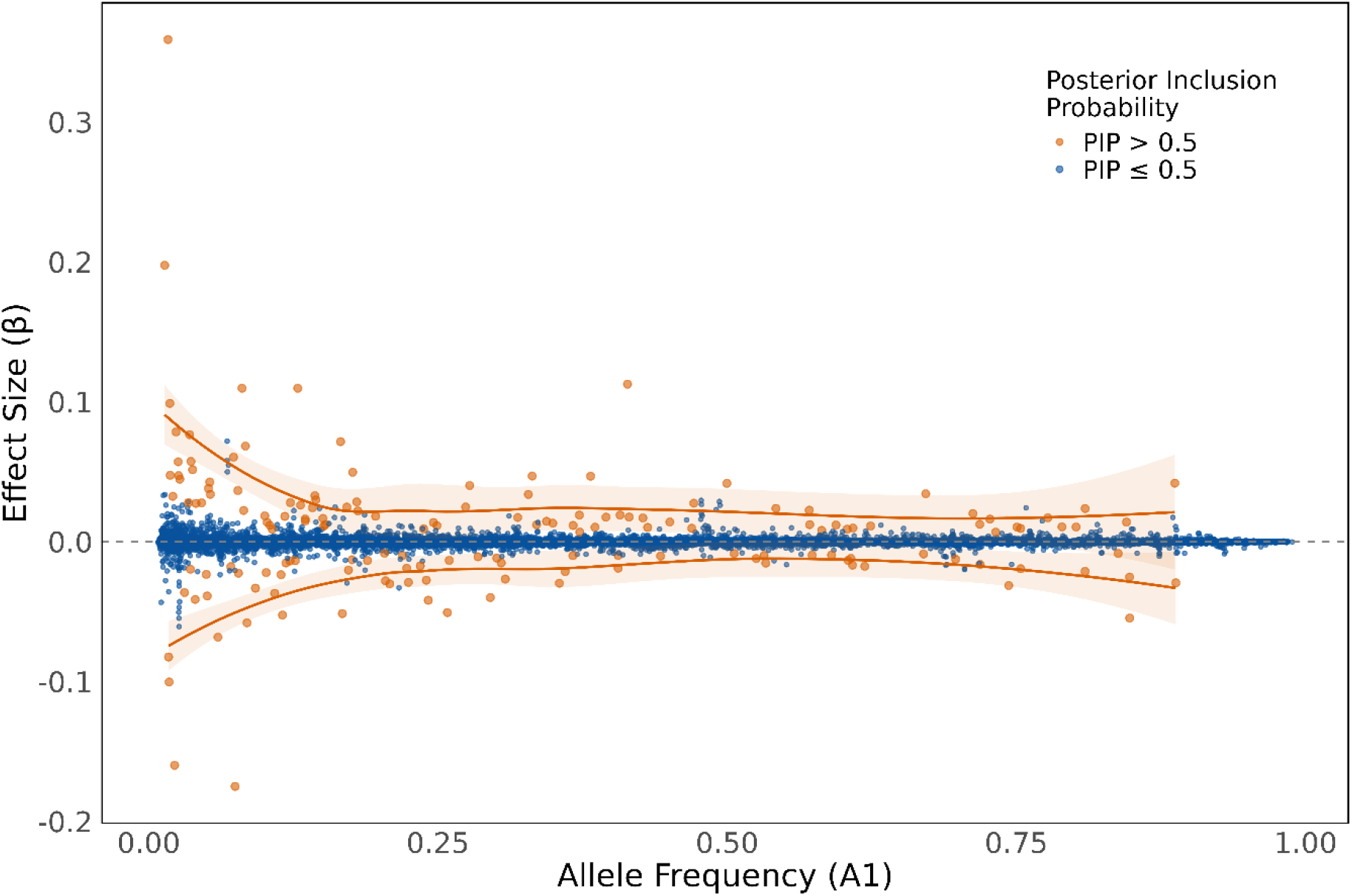
**Genetic effects across allele frequencies**, demonstrating an inverse relationship between effect size and frequency of the effect allele (n = 1.5M SNPs). It highlights the ability of SBayesRC to capture genetic signals across multiple ranges of allele frequencies and effect sizes. Findings demonstrated greater effect sizes in more rare genetic variants, a signature of natural selection. Moderate-confidence causal SNPs (PIP > 0.5) are illustrated in orange with their corresponding tendency of effect sizes across allele frequencies (orange line).

### Prioritization of Candidate Causal Genes

Functionality-informed genome-wide fine mapping assigned 17,150 SNPs to 581 high-confidence LCSs, each with 90% probability of containing at least one causal genetic variant (Figure 4). The average LCS contained 30 SNPs with the vast majority being multi-SNP sets (n = 544). A few high-confidence SNPs were assigned to their own LCS (n = 37). Proportion of the total genetic variance explained by the 17,150 prioritized SNPs in LCSs was estimated to be 35%.

**Figure 4:**
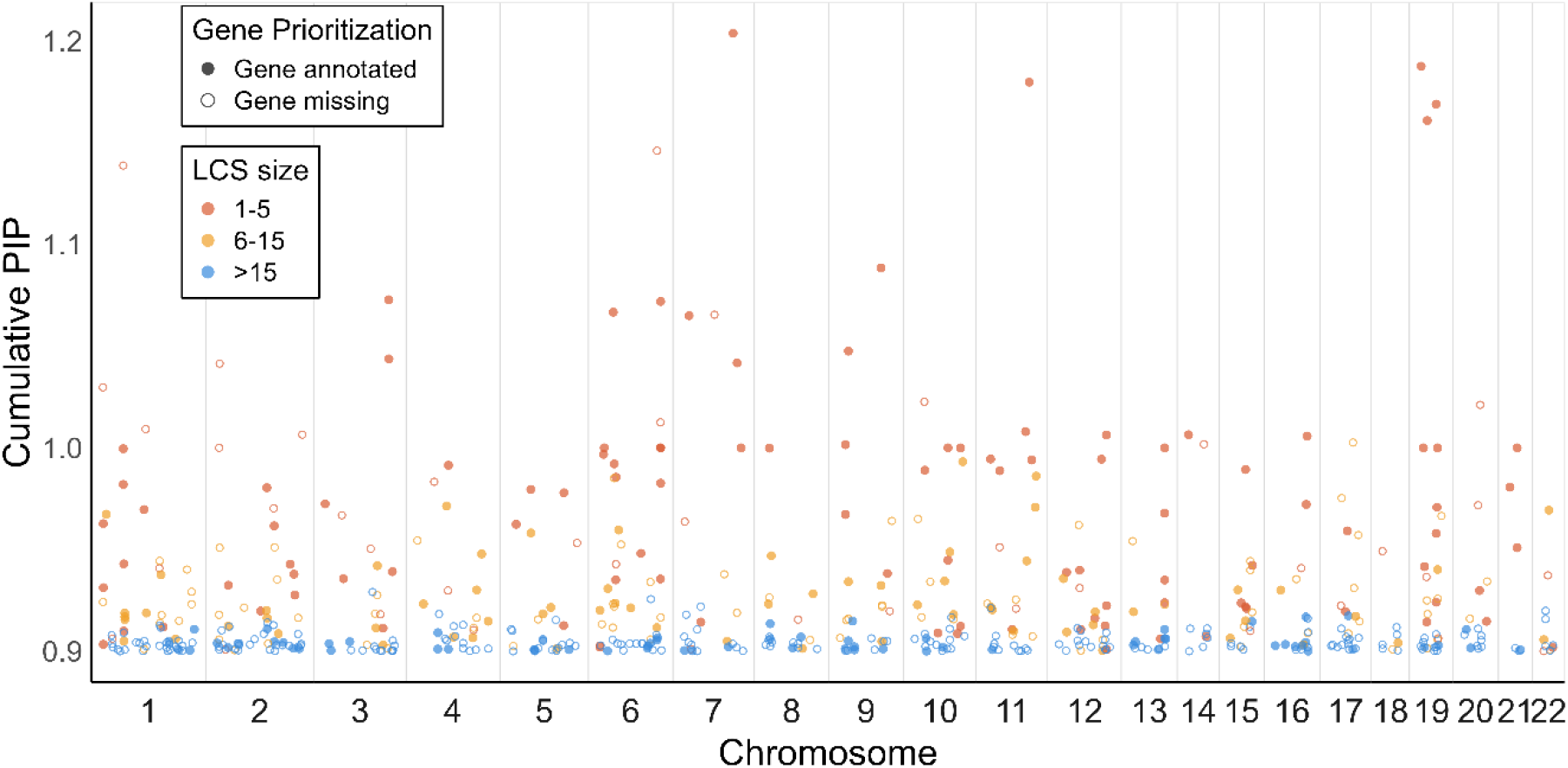
High-confidence credible sets and their cumulative PIP across chromosomes. 17,150 SNPs were assigned to 581 high-confidence LCSs (>90% probability of containing at least one causal genetic variant). 195 (34%) of LCSs were annotated to genes in a 5kB flanking window. Points are colored by LCS size, defined as the number of SNPs included in each set. High-resolution sets (1-5 SNPs) are shown in orange, moderate-resolution sets (6-15 SNPs) in yellow, and low-resolution sets (>15 SNPs) in blue. Gene-mapped sets (filled circles) are distinguished from sets with no gene annotation (hollow circles).

Nearest gene was prioritized in a 5kB flanking window to each of the high-confidence credible sets. This resulted in 195/581 (34%) LCSs to be mapped to genes of which 170/195 (87%) were recognized in the STRING database.

Annotated influential genetic variants contributing strongest to SNP-based heritability were linked to genes involved in processes affecting cardiovascular health and pathogenesis (Table 4). The risk variant that explained most genetic variance was mapped to the gene for phosphatase and actin regulator 1 (*PHACTR1*), playing a role in endothelial cell function and vascular homeostasis. The risk variant contributing second highest to SNP-based heritability but with highest individual effect was mapped to the gene for lipoprotein lipase (*LPL*), a key enzyme in triglyceride metabolism and clearance of circulating lipoprotein particles.

**Table 4:** Overview of genetic variants contributing strongest to genetic variance (Var_g_).

| SNP | Chr | Position | A1 | Frequency | $\beta$ | $\text{Var}_g$ | PIP | Gene |
| --- | --- | --- | --- | --- | --- | --- | --- | --- |
| rs9349379 | 6 | 12903957 | G | 0.41 | 0.11 | 2.49% | 1 | *PHACTR1* |
| rs7412 | 19 | 45412079 | T | 0.07 | - 0.17 | 1.59% | 1 | *APOE* |
| rs140570886 | 6 | 161013013 | C | 0.02 | 0.36 | 1.46% | 1 | *LPL* |
| rs28451064 | 21 | 35593827 | A | 0.13 | 0.11 | 1.04% | 1 | - |
| rs3918226 | 7 | 150690176 | T | 0.08 | 0.11 | 0.67% | 1 | *NOS3* |
| rs56393506 | 6 | 161089307 | T | 0.17 | 0.07 | 0.55% | 1 | *LPA* |
| rs1333046 | 9 | 22124123 | A | 0.50 | 0.03 | 0.46% | 0.2 | *CDKN2B-AS1* |
| rs186696265 | 6 | 161111700 | T | 0.01 | 0.20 | 0.42% | 0.9 | *LPA* |
| rs2266008 | 10 | 91014061 | G | 0.38 | 0.05 | 0.39% | 1 | *LIPA* |
| rs1333048 | 9 | 22125347 | C | 0.50 | 0.03 | 0.39% | 0.2 | *CDKN2B-AS1* |
| rs10455872 | 6 | 161010118 | G | 0.07 | 0.07 | 0.39% | 0.3 | *LPA* |
| rs9515203 | 13 | 111049623 | C | 0.26 | - 0.05 | 0.39% | 1 | *COL4A2* |
| rs9337951 | 10 | 30317073 | A | 0.33 | 0.05 | 0.38% | 1 | *JCAD* |
| rs116843064 | 19 | 8429323 | A | 0.02 | - 0.16 | 0.37% | 1 | *ANGPTL4* |
| rs10422256 | 19 | 11216617 | A | 0.50 | 0.04 | 0.36% | 0.9 | *LDLR* |
| rs10757279 | 9 | 22124630 | G | 0.48 | 0.03 | 0.35% | 0.3 | *CDKN2B-AS1* |
| rs2327429 | 6 | 134209837 | C | 0.30 | - 0.04 | 0.33% | 0.6 | - |
| rs2107595 | 7 | 19049388 | A | 0.18 | 0.05 | 0.32% | 0.9 | - |
| rs964184 | 11 | 116648917 | C | 0.85 | - 0.05 | 0.31% | 1 | *ZPR1* |
| rs74617384 | 6 | 160997118 | T | 0.07 | 0.06 | 0.31% | 0.3 | *LPA* |

The protective variant with strongest contribution to SNP-based heritability was mapped to the gene for Apolipoprotein E (*APOE*), a key component of circulating lipoprotein particles involved in lipid transport and clearance.

Three variants among the strongest contributors to genetic variance were not annotated to any gene. The strongest, rs28451064, explained 1.04% of genetic variation with high confidence of causality (PIP= 1).

### Protein-protein Interaction Network and Cluster Analysis

The protein-protein interaction network was significantly enriched with 78/170 (46%) proteins interacting, indicating proteins to be connected in specific biological processes associated with CAD (P = 2.22 × 10^−15^).

Cluster analysis resulted in the construction of three major clusters (Table 5 and Figure 5). The identified three major clusters showed interactive mechanisms via six bottleneck genes. The largest major cluster (n = 18 proteins) was related to vascular homeostasis. The second largest (n = 15 proteins) was related to lipoprotein function and cholesterol metabolism. The final major cluster (n = 12 proteins) was related to the AGE-RAGE signaling pathway. A total of 11 hub genes (≥ 5 interactions) were identified within the protein network as key regulators in CAD development. FN1, APOE, and PLCG2 were identified as the most interactive hub genes in each of the three major clusters respectively.

**Table 5:**
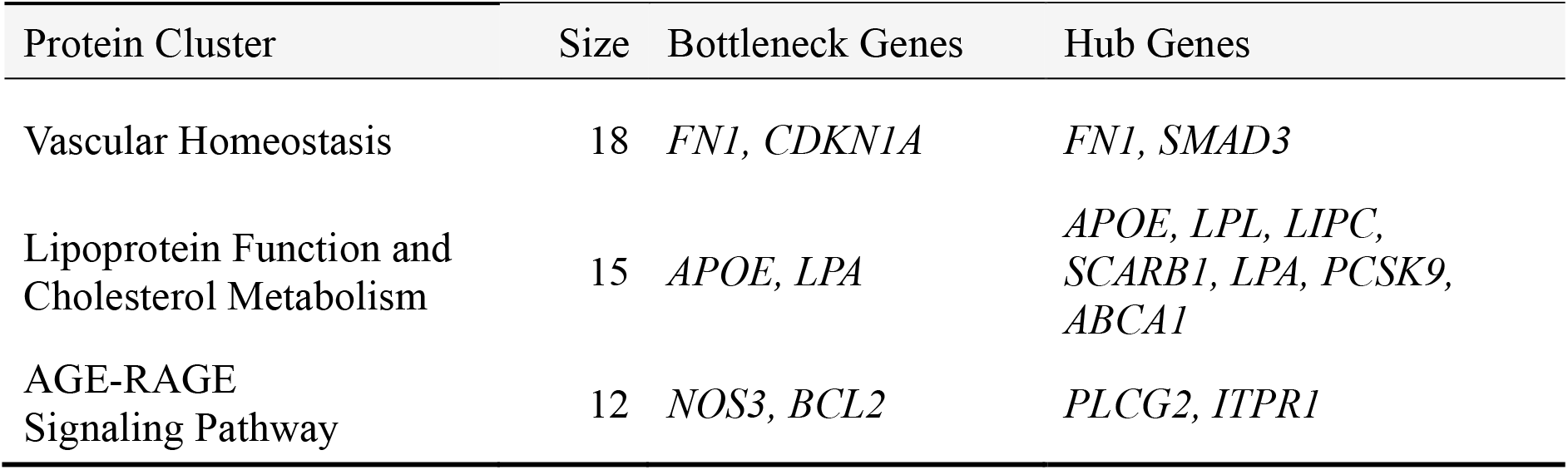
Major protein clusters representing dysregulated biological processes that drive genetic risk in CAD.

| Protein Cluster | Size | Bottleneck Genes | Hub Genes |
| --- | --- | --- | --- |
| Vascular Homeostasis | 18 | *FN1*, *CDKN1A* | *FN1*, *SMAD3* |
| Lipoprotein Function and Cholesterol Metabolism | 15 | *APOE*, *LPA* | *APOE*, *LPL*, *LIPC*, *SCARB1*, *LPA*, *PCSK9*, *ABCA1* |
| AGE-RAGE Signaling Pathway | 12 | *NOS3*, *BCL2* | *PLCG2*, *ITPR1* |

**Figure 5:**
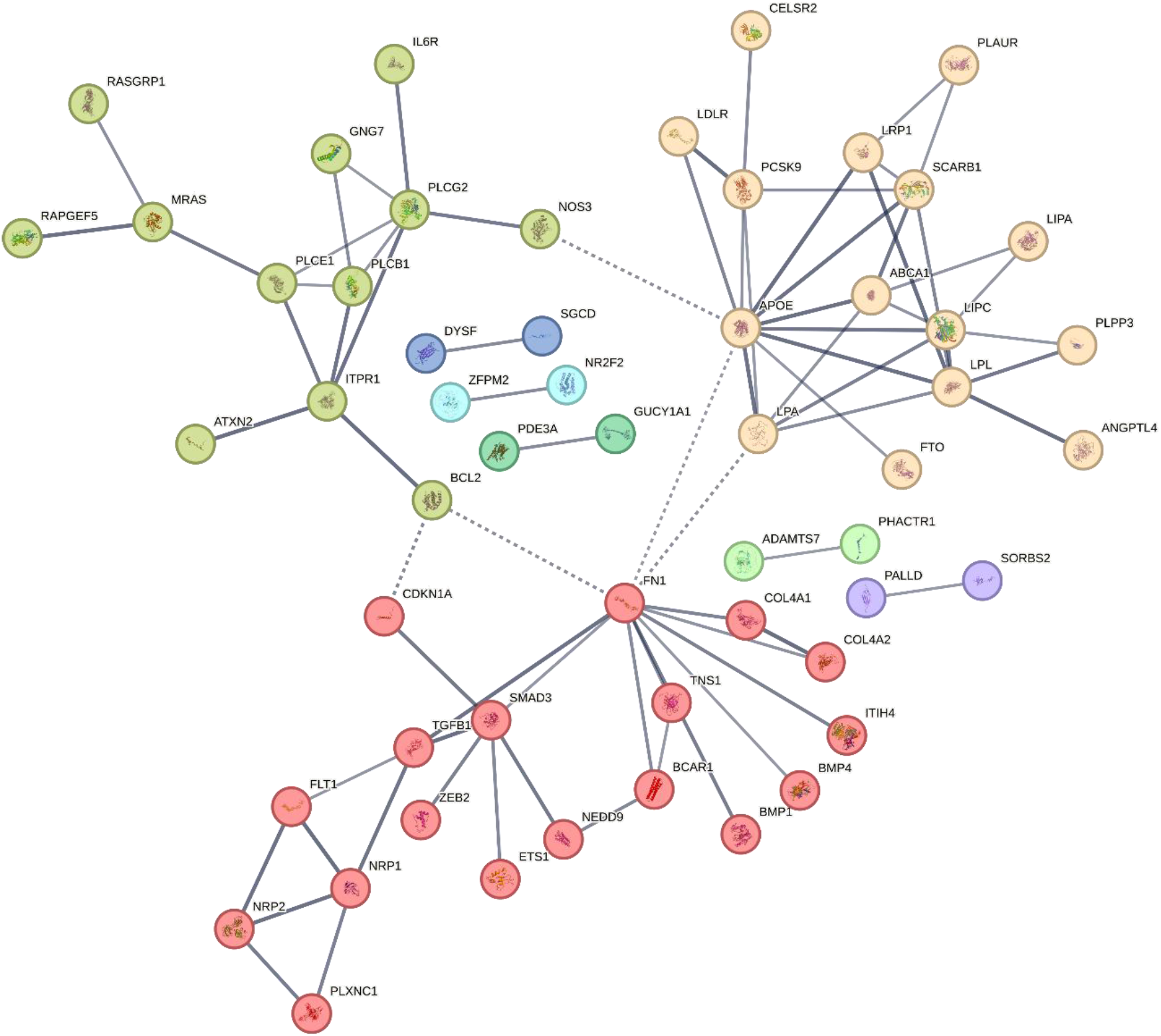
Protein-protein interaction network. Proteins were significantly interconnected (78/170 connected proteins), revealing the protein network to be enriched in central biological processes related to CAD pathogenesis. Three major clusters were identified, suggesting CAD pathogenesis to be driven by dysregulation in three distinct pathways: 1) Vascular homeostasis (red), 2) Lipoprotein function and cholesterol metabolism (beige), and 3) AGE-RAGE signaling pathway (green). Analysis revealed key regulators as hub genes with multiple interactions within each cluster. Bottleneck genes that interact with other major clusters are presented with dashed lines, while strength of interaction evidence is represented by line thickness with higher evidence having thicker lines. Each protein is labeled with their StringID from the STRING database.

### Pathway Enrichment Analysis

Pathway enrichment analysis on molecular level based on the Reactome database showed significant enrichment of prioritized genes in 16 pathways predominantly relating to lipid metabolism, vascular homeostasis and inflammatory response (Figure 6a). Lipoprotein assembly, modeling, and clearance were identified as the strongest association on molecular level.

**Figure 6:**
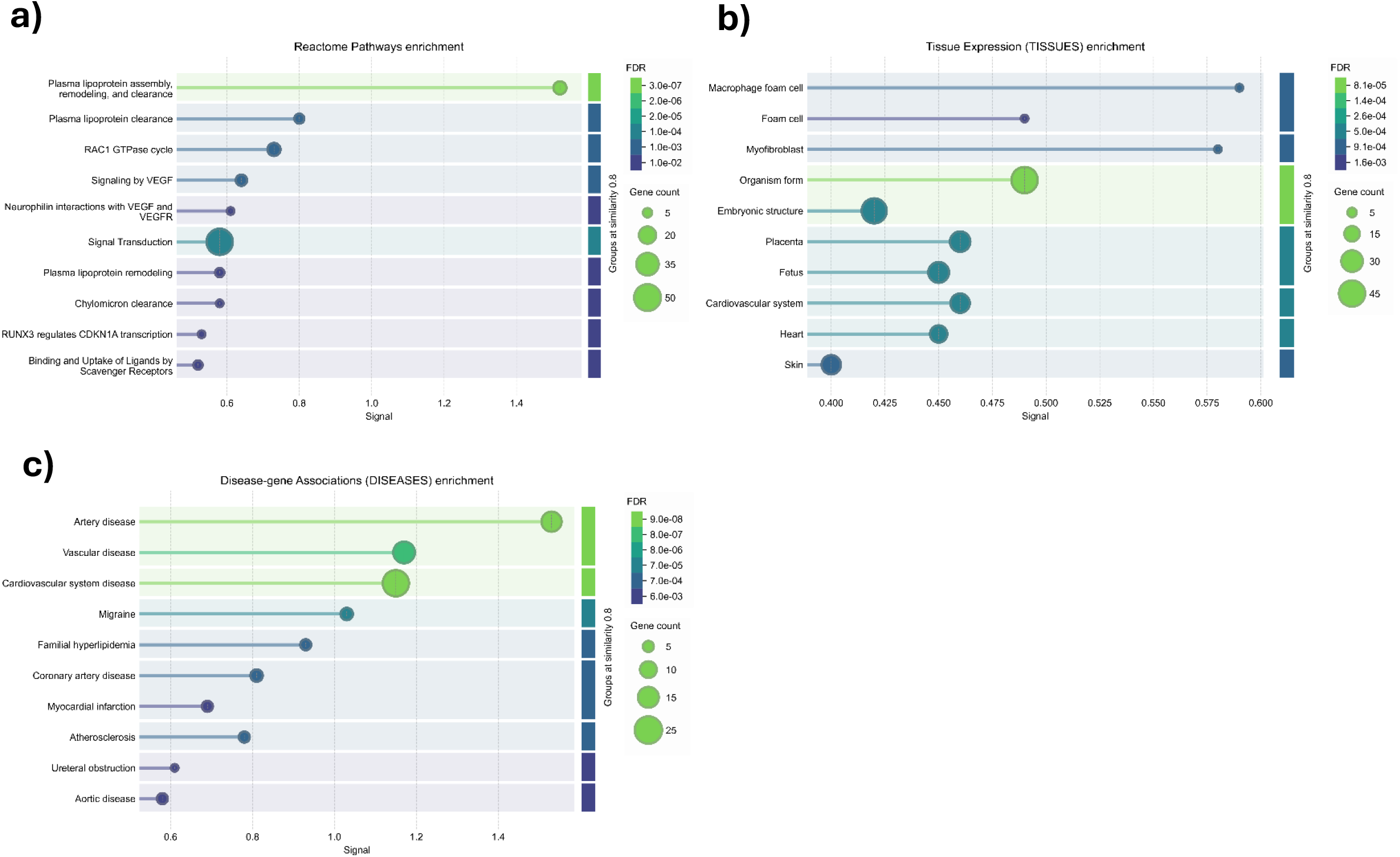
**Pathway enrichment analyses** of prioritized genes (n = 170 genes) compared with a random background gene set. **a) Molecular-level enrichment** based on terms related to biological pathways. Results showed significant enrichment predominantly related to lipoprotein function, endothelial homeostasis, and inflammatory responses. **b) Tissue-specific enrichment** based on tissue expression levels. Results demonstrated significant enrichment in key tissues relevant in CAD pathogenesis, identifying the macrophage foam cell as the highest associated term. **c) Disease-level enrichment** based on disease-associated terms. Results demonstrated significant enrichment in relevant diseases including cardiovascular diseases and other phenotypes overlapping with CAD. Each term is presented as circles, the size representing their included gene count, grouped by term similarity, and ranked by enrichment signal.

Enrichment analysis on tissue level based on the TISSUES database showed significant enrichment of prioritized genes in 21 tissues predominantly being highly vascularized or relating to inflammatory response or tissue regeneration (Figure 6b). Macrophage foam cells were identified as the highest-ranking contributor to CAD pathogenesis on the tissue level.

Enrichment analysis on disease level based on the DisGeNET database showed significant enrichment of prioritized genes in 17 diseases predominantly related to cardiovascular health with artery disease showing the strongest association (Figure 6c). Thus, validating the prioritized genes to associate with CAD and overlapping phenotypes.

## Discussion

### Functionality-informed Genome-wide Fine Mapping

The present study captured the highly polygenic architecture of CAD, on average estimating 34,468 genetic variants as causal associations explaining 3.8% of total phenotypic variance by SNP-based heritability.

Previous GWAS have reported approximately 400 independent genetic loci to be associated with CAD, while twin studies and updated genome-wide approaches have demonstrated approximately 40-60% heritability in CAD [8, 18, 19]. Bjorkegren et al reviewed the importance of the discovered significant loci in CAD, reporting that currently known genetic variants only approximately explain 10% of the total phenotypic variation in CAD [20]. The missing heritability in cardiovascular disease corresponds with the low estimated SNP-based heritability in the present study. This could possibly be explained by epigenetic factors, exclusion of rare variants (MAF < 0.01) and non-SNP variants (insertions and deletions), all contributing to heritability but not characterized in the present study [20, 21]. Rocheleau et al. evaluated the contribution on heritability of rare genetic variants using whole genome sequencing, estimating rare variants (MAF < 0.01) to explain approximately 50% of CAD heritability [22].

In the present study, 36 high-confidence SNPs (PIP > 0.9) were identified. 17,150 genetic variants in 581 high confidence credible sets were prioritized, linking them to 195 candidate genes influencing CAD risk and pathogenesis. Thus, 66% of the constructed sets were not annotated to any gene. This represents the huge challenge in transitioning from identifying the causal (rather than associated) variant to understanding the genetic influence on downstream biological processes that ultimately promote development of CAD. A narrow flanking window of 5kB was implemented to capture regulatory influence of variants in high resolution. The vast majority (> 90%) of the identified CAD-associated SNPs in GWAS are located in non-coding regions [23]. The influence on biological function of the majority of these variants is still unsettled, possibly acting over genomic distances of several hundred kilobases [7, 10]. This highlights the need for functional characterization of identified variants to move beyond statistical association toward mechanistic understanding of their role in CAD pathogenesis.

In the present study, functional annotations were implemented in a Bayesian approach using state of the art SBayesRC to model the functional association between genotype and phenotype. The current findings highlight the value of a Bayesian fine mapping approach that accounts for LD structure and incorporate functional annotations to apply appropriate effect size shrinkage and prediction of candidates for causal associations. This functionality-informed approach has previously been demonstrated to provide new insights into the genetic architecture of multiple complex polygenic traits. Enriched fine mapping with functional annotations has been demonstrated to increase the number of estimated high-confidence associations and improve mapping calibration, resulting in enhanced polygenic prediction and variant prioritization [14, 24, 25, 26]. Zeng et al. demonstrated SBayesRC to outperform existing fine-mapping techniques (FINEMAP, SuSiE, FINEMAP-Inf, and SuSiE-Inf) in mapping precision and prediction accuracy [12]. They likewise demonstrated that SBayesRC yielded robust polygenic predictions, outperforming other methods such as LDpred2, LDpred-funct, MegaPRS, PolyPred-S, and PRS-CSx [11].

SBayesRC was performed based on GWAS summary statistics and LD reference data from individuals with predominantly European ancestry. While ancestral homogeneity improves model convergence and LD consistency, it limits the generalizability of the estimated genetic contributions to non-European populations.Future studies incorporating multi-ancestry GWAS summary statistics and LD data will therefore be essential to evaluate whether the identified variants exert similar effects across diverse populations.

With the arrival of increasingly more powerful GWAS, increasing evidence supports the hypothesis of genome-wide signatures of natural selection on genetic variants associated with complex polygenic traits such as CAD [27]. Zeng et al. performed Bayesian modeling with MAF as a parameter (SBayesS) and demonstrated strong signals of natural selection on variants in multiple polygenic traits including cardiovascular disease. They demonstrated genetic variants with deleterious effects to have larger effect sizes and be kept at lower frequencies by negative selection and having higher probabilities to be in functional genetic regions [28]. In the present study, greater effect sizes in more rare genetic variants were observed, demonstrating the same signature of natural selection in CAD.

### Genetic Contribution in CAD Pathogenesis

The present study identified multiple high-confidence genetic polymorphisms that individually contributed substantially to SNP-based heritability in CAD. Overall, they were all linked to genes influencing key functions in cardiovascular homeostasis. Analyses of the prioritized genes in protein-protein interaction networks and pathway enrichment highlighted their critical biological influence in disease-associated pathways. 78 protein encoding genes with strong evidence of interaction were identified based on the STRING database, functioning in three major protein clusters relating to key disease mechanisms. These findings build on existing evidence, highlighting well established risk factors such as altered lipid metabolism while also suggesting underlying molecular and cellular mechanisms influencing vascular function and inflammatory responses to drive CAD pathogenesis (Figure 7).

**Figure 7:**
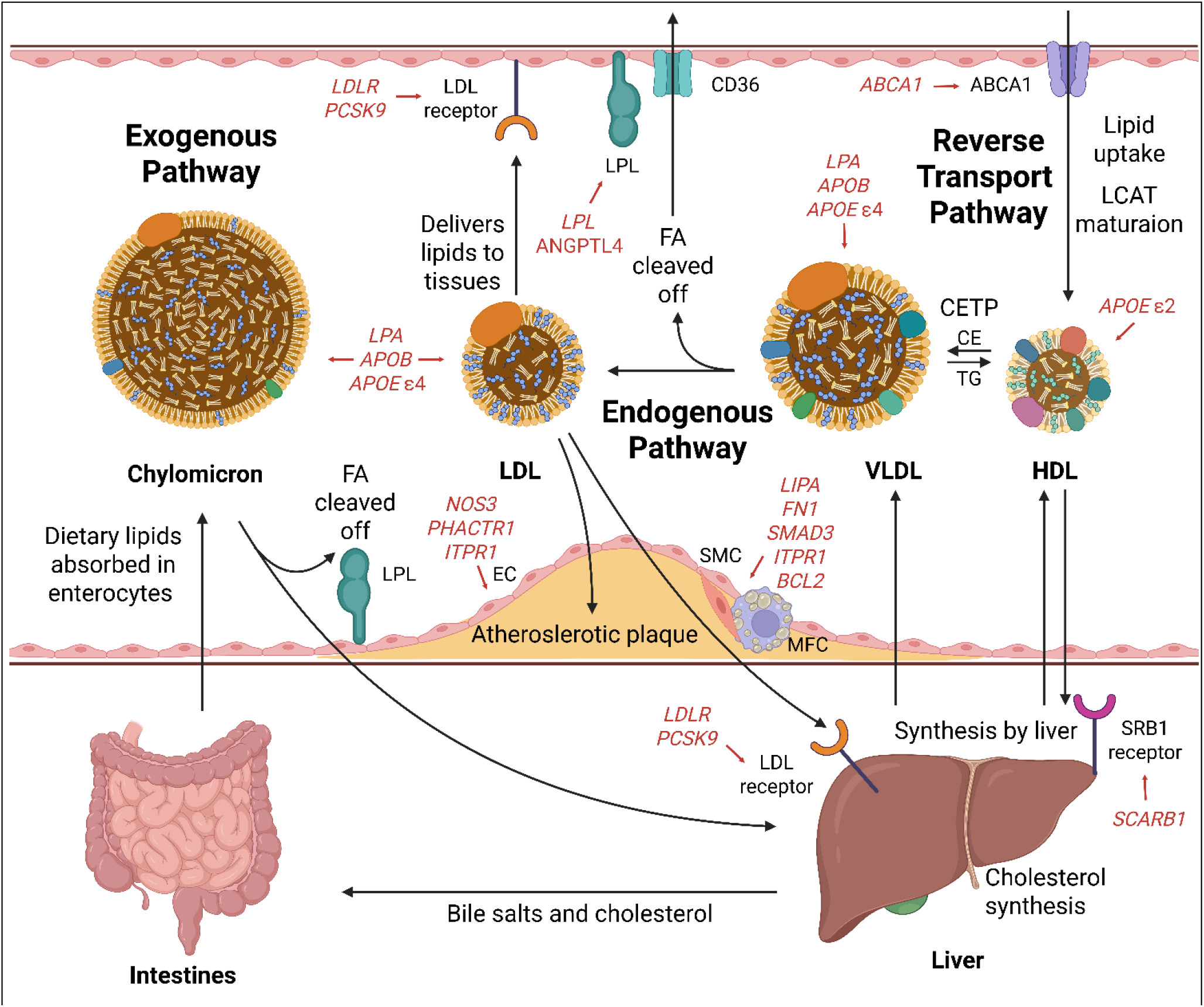
Genetic contribution in CAD pathogenesis. The prioritized genetic variants converged on three interlinked biological pathways: 1) lipoprotein function and cholesterol metabolism, 2) vascular homeostasis, and 3) cellular stress responses and inflammation. Highlights the genetic influence of the most influential variants on biological processes in CAD pathogenesis.

### Lipoprotein Function and Cholesterol Metabolism

The current study demonstrated CAD risk to be driven by impaired lipoprotein function and cholesterol metabolism. These findings build on existing evidence that genetic accumulation of risk variants lead to increased CAD susceptibility through altered cholesterol metabolism and lipoprotein assembly, remodeling, and clearance that lead to dyslipidemia [29]. Comprehensive amounts of evidence in literature demonstrate impaired lipid metabolism as one of the strongest risk factors in CAD. Ajoolabady et al. reviewed the pathogenic mechanisms in atherosclerosis, highlighting LDL-cholesterol as the predominant lipid component during build-up of inflammatory plagues in the arterial walls. In the plaques, oxidized LDL cholesterol promotes uptake by macrophages that differentiate into proinflammatory foam cells that drive inflammatory responses, resulting in atherosclerotic progression, vascular inflammation, and arterial stenosis [30].

In the present study, multiple influential variations were identified to affect circulating lipoprotein particle function. Some of the strongest influential variants were linked to genes encoding lipoprotein(a) (LPA) and apolipoprotein E *ϵ*2 isoform (APOE), both demonstrated as strong predictors to cardiovascular disease. LPA consists of a lipid carrying LDL-like core linked to apolipoprotein(a). LPA is demonstrated to inhibit plasminogen and to function as carriers of pro-inflammatory oxidized phospholipids, while also showing increased affinity to endothelial cells, macrophages, and platelets [31, 32]. In contrast, APOE *ϵ*2 has been associated with increased human longevity, demonstrating protective functions in cardiovascular disease caused by altered lipoprotein receptor affinities and modulations on inflammatory processes [33, 34]. Genetic variants influencing these genes alter the atherogenic properties of circulating lipoprotein particles and affect vascular homeostasis.

Multiple influential variants were linked to genes that influence key pathways of lipoprotein homeostasis. This includes genes encoding LDL receptor (LDLR), proprotein convertase subtilisin/kexin 9 (PCSK9), lipoprotein lipase (LPL), angiopoietin-like 4 (ANGPTL4), and zinc finger protein ZPR1 (ZNF259). LDLR facilitates clearance of circulating LDL particles by receptor-mediated endocytosis in the liver [35], while PCSK9 induces degradation of LDLR [36]. LPL located in the vascular wall hydrolyzes triglycerides from circulating triglyceride-rich VLDL particles [37] and is inhibited by ANGPTL4 [38]. ZNF259 is a regulatory protein that interacts with the gene encoding APOA5, which activates LPL [39]. Dysfunction of LDLR, PCSK9, LPL, ANGPTL4, and ZNF259 thus leads to altered remodeling of circulating lipoprotein particles. Dysregulation in these pathways would thus cause increased levels of atherogenic lipoprotein particles in the blood, accumulation of lipids in the vascular wall, and acceleration of plaque build-up. Further, the identified association between genes influencing LPL function and CAD risk provides genetic evidence supporting the emerging causal role of triglyceride-rich lipoprotein particles in CAD pathogenesis.

Other influential variants were linked to genes encoding ATP-binding cassette transporter A1 (ABCA1), hepatic lipase (LIPC), and scavenger receptor class B member 1 (SR-BI). ABCA1 interacts with HDL particles and promotes the efflux of excess intracellular lipids in the peripheral tissues that load HDL particles with cholesterol and phospholipids [40]. LIPC does not only hydrolyze triglycerides in VLDL particles, it also effectively remodels HDL particles and thus affects levels of HDL-C [41]. SR-BI is a major receptor for HDL that facilitates selective uptake of cholesterol from HDL in the liver [42]. They all play key roles in the reverse cholesterol transport pathway that clears excess cholesterol from the peripheral tissues and transports it to the liver.Dysregulation in pathways affecting HDL homeostasis would disrupt cholesterol efflux capacity and reduce the clearance of atherogenic lipoprotein particles [43, 44]. Further, HDL particles mediate cardiovascular-protective functions beyond those of cholesterol metabolism. This includes antioxidant, anti-inflammatory, and antithrombotic capacities, which modulate vascular homeostasis. An atherogenic shift in HDL function thus contributes to systemic vascular inflammation and endothelial dysfunction, accelerating atherogenesis [43, 44].

Finally, one influential variant was linked to the gene encoding lysosomal acid lipase (LIPA). LIPA is responsible for the lysosomal degradation of cholesteryl esters and triglycerides that are taken up by macrophages and invading smooth muscle cells in the vascular wall [45]. Impaired LIPA function leads to disruption of intracellular lipid metabolism, accumulation of cytosolic lipid droplets, and promotion of pro-inflammatory foam-cell cellular phenotypes that drive vascular inflammation and plaque build-up [46].

### Vascular Homeostasis

This study builds on the hypothesis that disruption in vascular homeostasis not only functions as a marker of CAD but rather as a major driver in CAD pathogenesis through accumulation of genetic risk variants that ncrease susceptibility to CAD. The importance of endothelial function in CAD is evident in the literature, reporting the presence of endothelial dysfunction as one of the first recognizable signs in early atherosclerosis and multiple other age-related diseases [47]. Likewise, vascular smooth muscle cells play a major role in all stages of atherosclerosis. Invading smooth muscle cells undergo metaplasia and shift into proliferative, synthetic, or even foam-cell-like phenotypes that contribute to plaque build-up [48]. Finally, the extracellular matrix demonstrates an active role in CAD pathogenesis by dynamically controlling structural characteristics and cellular signaling pathways in the vascular microenvironment [49].

The present study identified multiple influential variants affecting genes that modulate vascular homeostasis via intracellular signaling pathways. This includes genes encoding phosphatase and actin regulator 1 (PHACTR1), cyclin dependent kinase inhibitor 1A (CDKN1A), and SMAD family member 3 (SMAD3). PHACTR1 plays a key role in regulation of cell cytoskeleton dynamics, motility and survival, and has been demonstrated to associate with proatherogenic conditions such as oxidation of LDL, proinflammatory processes, and reduced nitric oxide production [50]. CDKN1A regulates cell-cycle processes and has been associated with CAD possibly through promotion of endothelial cell senescence and vascular inflammation [51]. SMAD3 acts as a key transcriptional regulator within the transforming growth factor-beta (TGF-β) signaling pathway. It modulates pivotal signaling pathways important for cell differentiation and proliferation, immune responses, tissue regeneration, and fibrosis [52].

Other variants were linked to genes acting in pathways regulating extracellular signaling pathways. This includes genes encoding alpha-2 chain of type IV collagen (COL4A2), junctional cadherin 5-associated protein (JCAD), and fibronectin 1 (FN1). COL4A2 functions as a structural component in collagen IV, crucial for basement membrane integrity and functionality. It underlies the endothelial cells and surrounds the vascular smooth muscle cells, serving both as extracellular scaffold and modulator of cellular functions [53]. JCAD is an endothelial cell-cell junction, regulating vascular integrity by acting on coagulation, fibrinolysis, sheer stress responses, barrier function, angiogenesis, and vascular remodeling [54, 55]. FN1 acts as crucial structural component and modulator of the extracellular matrix assembly. It modulates vascular tone, vascular remodeling, inflammatory processes, and cell proliferation [56].

Findings in the present study thus demonstrate genetic risk variants to influence vascular homeostasis that disrupt critical cellular functions, promote vascular inflammation, alter the extracellular environment, and impair vascular remodeling. Consistent with this, Gutierrez et al. highlights that impaired vascular function in nutrient trafficking, tissue repair, blood vessel homeostasis, and blood flow would result in reduced ability to handle cellular stress and inflammatory processes. This ultimately results in promotion of atherogenic vascular remodeling that drives CAD pathogenesis [57].

### AGE-RAGE Signaling Pathway and Inflammation

Lastly, the present study identified CAD risk to be driven by altered functions in the AGE-RAGE signaling pathway, a key modulator of inflammatory responses. Large amounts of evidence link atherosclerosis closely with chronic inflammation in the arterial walls, and previous studies have shown that the AGE-RAGE signaling pathway initiate and accelerate plaque formation, even independently of diabetes. Advanced glycation end products (AGE) are formed in tissues during natural aging processes, with accumulation being associated with multiple human diseases including CAD. The receptor of advanced glycation end products (RAGE) initiates production of reactive oxygen species and recruitment of proinflammatory cells [58]. Results from the present study propose that CAD susceptibility is driven by accumulation of genetic risk variants disturbing the homeostasis in the AGE-RAGE signaling pathway. Consistent with these results, Wang et al. reviewed that excessive activation of the AGE-RAGE pathway led to vascular inflammation, promotion of oxidative stress, endothelial dysfunction, impairment of cholesterol clearance, and arterial damage, resulting in accelerated formation of atherosclerotic plaques [58].

The present study identified multiple influential variants that were linked to genes acting in pathways of inflammatory processes. This includes genes encoding endothelial nitric oxide synthase 3 (NOS3), B-cell lymphoma 2 (BCL2), phospholipase C-gamma-2 (PLCG2), and inositol 1,4,5-trisphosphate receptor type 1 (IP3R1). NOS3 synthesizes nitric oxide, which has been demonstrated to have cardiovascular-protective effects. Besides modulating vascular tone, nitric oxide regulates endothelial cell survival, reduces oxidation of LDL-C, and inhibits vascular wall adhesion of platelets, immune cells, and smooth muscle cells [59]. BCL2 plays a critical role in anti-apoptotic processes. Downregulation of BCL2 in advanced atherosclerotic plaques indicates hyperactivation of apoptotic signals [60]. Promotion of vascular cell survival, possibly via the NOS3 signaling pathway, is suggested as functions of BCL2, however the underlying mechanistic effects in cardiovascular disease are not yet understood [61]. The present study connects BCL2 function in CAD pathogenesis with pathways of vascular homeostasis, demonstrating interactions with CDKN1A and FN1 in the protein-protein interaction network. PLCG2 is an essential regulator of inflammatory response and immune cell infiltration, contributing to vascular inflammation in cardiovascular disease [62]. IP3R1 plays a central role in intracellular calcium signaling. This pathway modulates vascular tone [63], affects intracellular stress of smooth muscle cells, endothelial cells, and immune cells [64], and possibly impact LDL-particle remodeling via PCSK9 expression [65].

The present study thus contributes with genetic evidence that dysregulation in pathways of inflammatory processes drives CAD pathogenesis, even linking it with key pathways influencing vascular homeostasis and lipid metabolism.

Furthermore, multiple identified variants with high genetic contribution but lower estimated causal probability were linked to the gene CDKN2B-AS1 in the 9p21 CAD risk locus. This risk locus has been demonstrated to play critical roles in age-related diseases by modulating cellular senescence, inflammatory pathways, cell metabolism, and vascular aging [66].

Finally, three variants (rs28451064, rs2327429, and rs2107595), were not annotated to any gene, despite major contribution to heritability and moderate to high confidence of causality (PIP = 1, 0.6, and 0.9 respectively). Knowledge of variant rs28451064 and its influence on genetic function and association with cardiovascular disease is very sparse. Previous studies have also identified the genetic region as a CAD risk locus. The rs28451064 variant is suggested to influence mitochondrial function and cellular stress responses, possibly leading to endothelial dysfunction and increased CAD susceptibility [67, 68]. Likewise, the underlying biological mechanism of the variant rs2327429 is unclear. The variant is suggested to influence smooth muscle cell function and ascular remodeling [69]. Previous studies have associated the variant rs2107595 with atherosclerosis and cardiovascular disease, possibly via the epigenetic regulator HDAC9 that affects inflammatory processes, immune cell function, and cholesterol metabolism [70].

## Conclusion

This study applied functionality-informed genome-wide fine mapping to dissect the genetic basis of CAD. By modeling the contributions of 6.9 million common variants, the complex polygenic nature of CAD was confirmed. A small group of highly influential variants was identified, however with the vast majority of variants exerting small individual effects that collectively account for a substantial proportion of SNP-based heritability in CAD.

The prioritized candidates converged on three interlinked biological pathways: 1) lipoprotein function and cholesterol metabolism, 2) vascular homeostasis, and 3) cellular stress responses and inflammation, reflecting the multifaceted pathogenesis of CAD. Findings highlight how disruption of lipid homeostasis at multiple levels collectively contribute to CAD risk, consistent with the central role of dyslipidemia in atherosclerotic pathogenesis. Within vascular homeostasis, implicated genes point to dysregulation of endothelial integrity, smooth muscle cell phenotype, and extracellular matrix assembly as central mechanisms driving atherosclerotic risk. The intersection of inflammatory signaling to lipid metabolism and vascular dysfunction further highlights the interdependence of these pathogenic axes, consistent with the emerging understanding of CAD as a chronic inflammatory disease of the vessel wall.

A Bayesian functionality-informed approach represents a meaningful framework to understand the genetic basis in CAD, enabling the prioritization of biologically plausible causal variants rather than relying solely on statistical significance. Notably, a substantial proportion of prioritized variants were not mapped to genes, residing in intergenic regions of the genome. This highlights a critical gap in the current mechanistic understanding of CAD genetics as the regulatory functions of these variants remain largely uncharacterized. Future mechanistic studies combined with research integrating biological information in genetic modeling will be essential to elucidate the biological relevance of these variants and their contribution to CAD pathogenesis. As analyses were based on European ancestry cohorts, generalizability to diverse populations remains limited. Future multi-ancestry studies will be essential to fully characterize the global genetic landscape of CAD.

Collectively, these findings advance the mechanistic understanding of CAD pathogenesis and provide a foundation for future research in functional genetics, ultimately supporting the translation of genomic discoveries into therapeutic targets and personalized prevention strategies.

## Data Availability

All source data were openly available before the initiation of the study. All data analyzed in this study are derived from publicly available summary-level datasets. No individual-level data were accessed. All tools and databases utilized are freely available to the public. All data produced in the present study are available upon reasonable request to the authors.
The genome-wide association study (GWAS) summary statistics used in this study are publicly available from the original consortia and repositories. Linkage disequilibrium data and functional annotations (BaselineModel2.2) are publicly available as part of the original SBayesRC GitHub resource. The utilized gene map is publicly available as part of the original GCTB resource. Protein-protein interaction network construction and pathway enrichment analyses were carried out using publicly available resources accessible through the STRING database.

https://kp4cd.org/dataset_downloads/mi#summary

https://github.com/zhilizheng/SBayesRC

https://gctbhub.cloud.edu.au/software/gctb/#Genome-wideFine-mappinganalysis

https://string-db.org/

